# Prediction of Ciprofloxacin Resistance in Hospitalized Patients Using Machine Learning

**DOI:** 10.1101/2022.10.18.22281205

**Authors:** Igor Mintz, Michal Chowers, Uri Obolski

**Affiliations:** School of Public Health, Tel Aviv University, Tel Aviv, Israel; Porter School of the Environment and Earth Sciences, Tel Aviv University, Tel Aviv, Israel; Meir Medical Center, Kfar Saba, Israel; Sackler School of Medicine, Tel Aviv University, Tel Aviv, Israel

## Abstract

**Background:** Ciprofloxacin is a widely used antibiotic that has lost efficiency due to extensive resistance. We developed machine learning (ML) models that predict the probability of ciprofloxacin resistance in hospitalized patients.

**Methods:** Data were collected from electronic records of hospitalized patients with positive bacterial cultures, during 2016-2019. Susceptibility results to ciprofloxacin (n=10,053 cultures) were obtained for *E. coli, K. pneumoniae, M. morganii, P*.*aeruginosa, P. mirabilis* and *S. aureus*. An ensemble model, combining several base models, was developed to predict ciprofloxacin resistant cultures, either with (gnostic) or without (agnostic) information on the infecting bacterial species.

**Results:** The ensemble models’ predictions were well-calibrated, and yielded ROC-AUCs (area under the receiver operating characteristic curve) of 0.763 (95%CI 0.634-0.785) and 0.849 (95%CI 0.799-0.921) on independent test-sets for the agnostic and gnostic datasets, respectively. Shapley additive explanations analysis identified that influential variables were related to resistance of previous infections, where patients arrived from (hospital, nursing home, etc.), sex, and recent resistance frequencies in the hospital. A decision curve analysis revealed that implementing our models can be beneficial in a wide range of cost-benefits considerations of ciprofloxacin administration.

**Conclusions:** This study develops ML models to predict ciprofloxacin resistance in hospitalized patients. The models achieved high predictive ability, were well calibrated, had substantial net-benefit across a wide range of conditions, and relied on predictors consistent with the literature. This is a further step on the way to inclusion of ML decision support systems into clinical practice.

## Introduction

Antimicrobial resistance (AMR) has developed into a global public health crisis. AMR often emerges rapidly in bacterial populations, and the effectiveness of newly introduced antibiotics can substantially drop after a few years of clinical use (1, 2). In settings of high resistance levels, such as treatment of hospitalized patients, it may become challenging to find empiric antibiotic treatments which will be effective, while minimizing collateral resistance (3). Such inappropriate empirical treatment is associated with the prevalence of AMR (4). Despite guidelines (5), literature on collateral damage of antibiotics (5, 6), and stewardship initiatives (7), the frequency of bug-drug mismatch in empiric treatment often remains high (4, 8).

A notable example of a broadly used antibiotic, with increasing concerns about its resistance frequencies, is ciprofloxacin. Ciprofloxacin is a fluoroquinolone antibiotic, which has been widely used since the early 2000s and is currently on the World Health Organization’s List of Essential Medicines (9). Ciprofloxacin is effective against various gram-negative bacteria, and to a lesser extent gram-positive bacteria, and is used in the treatment of urinary tract, respiratory tract, bone and joint, intra-abdominal, and other infections (10, 11). Hence, ciprofloxacin has been the drug of choice for many infections both in in- and out-patient settings. High consumption rates over decades inevitably increased resistance to the drug (12–14), with an additional indirect effect on non-consumers (15), impeding effective therapy (16). However, reversion to high levels of sensitivity to quinolones is rapid upon decrease in quinolone consumption (17). Therefore, minimizing unnecessary ciprofloxacin use can have substantial public health impact.

The use of machine learning (ML) in the context of AMR has been rapidly increasing with the availability of electronic medical records (EMRs) and development of new algorithms. ML models are potentially nearing the point where they can support clinicians’ decisions of empiric therapy, by providing rapid predictions of resistance (18, 19). Hence, constant improvement of the methodology and outcomes of such models is of high importance. In the context of ciprofloxacin, prediction models have been scarce and limited to community-acquired urinary tract infections (20), only to intensive care units (21), specific site of infection (22), or to specific subsets of patients (23).

In this study, we developed an ensemble ML model that predicts resistance to ciprofloxacin based on hospitalized patients’ EMRs. Importantly, we include as variables relevant frequencies of resistance within the hospital, and not solely the examined patient’s EMR. Our models are applied to two settings: assuming that the infecting bacterial species is unknown (a bacteria-agnostic dataset) or known (the bacteria-gnostic dataset). Furthermore, explainability methods are used to analyze important predictors of resistance in our ML models

## Methods

### Data

Data were retrieved from Meir Medical Center, a hospital in Israel which serves approximately 600,000 residents. EMRs of patients who had positive bacterial cultures that were tested for ciprofloxacin susceptibility between the years 2016-2019 were retrieved. The data contained information regarding patients’ demographics, functional status, previous antibiotics usage and previous hospitalization within the previous year, bacterial pathogen and susceptibility results. Bacterial cultures demonstrating intermediate resistance results were regarded as resistant.

Additional features related to previous infections with resistant bacteria, previous antibiotic usage, and previous hospitalizations were engineered from the patients’ EMRs. The final dataset contained 10,053 susceptibility test results of 5,540 patients and 73 variables (see Supplementary Table S1). These data were used to create two data sets: bacteria gnostic (the whole data) and bacteria agnostic (without 20 features related to the bacteria). Each dataset was divided into a training set (75% of all samples) and a test set (25% of all samples), based on the date the culture was taken (Figure 1). All the presented results were obtained when training the models solely on the training set, and testing them on the independent test set.

**Figure 1:**
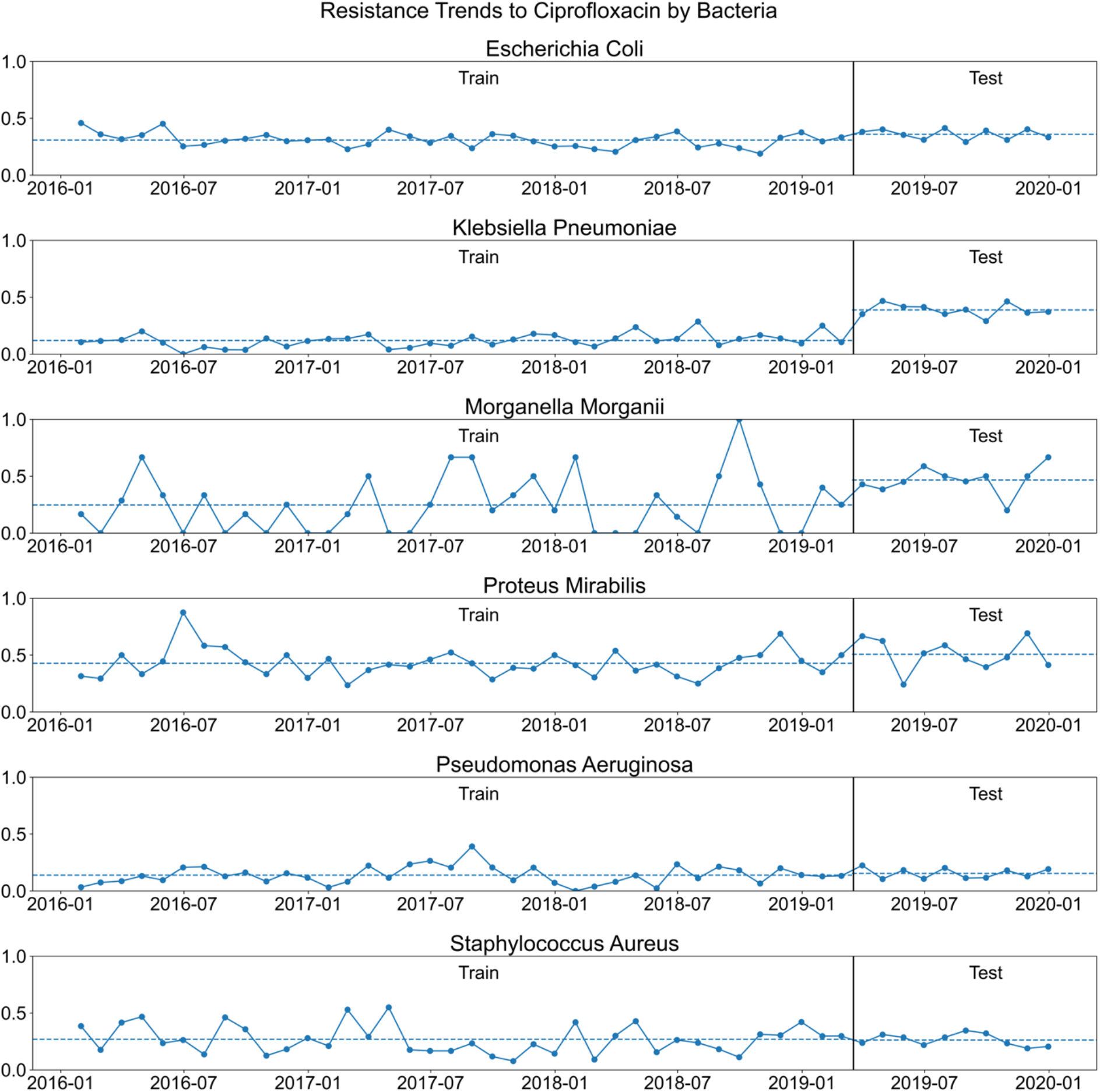
Ciprofloxacin resistance time-trends stratified by bacterial species. Points connected by solid lines are the average monthly ciprofloxacin resistance frequencies. The dotted horizontal lines represent the average resistance in the training and test sets, which are separated by the black vertical lines.

### Machine learning algorithms

We used an ensemble of several ML algorithms, which we term ‘base learners’: LASSO penalized logistic regression (24), random forest (25), gradient-boosted trees (25), and neural networks (25). The base learners’ hyperparameters were optimized using 200 random searches (26) with a five-fold, time series cross-validation. To improve the predictions of the four base learners, a stacking technique was applied. In this technique, the predictions of the base learners are given as inputs to a second-level learning algorithm (super learner). The super learner was a logistic regression algorithm trained to optimize the predictions (27). We adopted a process described elsewhere (28) to train the super learner on time series data (Figure S1 in the Supplementary Material). This resulted in a single ensemble model whose output is the predicted probability of the culture result to have resistance to ciprofloxacin. The tuned hyperparameters are shown at Supplementary Table S2. Model performance was evaluated using the area under the receiver operating characteristic curve (ROC-AUC) metric. Confidence intervals (CI) were calculated using 5,000 bootstrap samples of the test-set data. Model agnostic approximation of the Shapley additive explanations (SHAP) was performed with “kernel SHAP” (29), employing 300 background samples from the training data and calculating the SAHP values of the entire test set.

### Decision curve analysis

A decision (also known as a utility) curve analysis was performed using the predictions of our ensemble model on the test-set. In this analysis, the standardized net benefit (sNB) of a decision is defined by the following equation (30, 31):

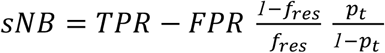

where TPR and FPR are the true- and false-positive rates, respectively; *p*_*t*_ is a threshold probability; and *f*_*res*_ is the frequency of resistant infections. In our case, *p*_*t*_ is the threshold probability above which a decision maker (i.e., clinician) is willing to act as if the infection is resistant to ciprofloxacin. This implies that the cost of falsely deciding that an infection is susceptible to ciprofloxacin is *p*_*t*_*/*(*1* − *p*_*t*_) fold the benefit of correctly deciding it is susceptible to ciprofloxacin. Hence *p*_*t*_*/*(*1* − *p*_*t*_) is also termed the cost-benefit ratio. For example, assume a clinician may not treat an infection with ciprofloxacin when she knows that the probability of ciprofloxacin resistance is above 0.2, but will treat them with ciprofloxacin otherwise. The clinician is hence implicitly willing to inefficiently treat one patient with a ciprofloxacin resistant infection for every four patients with susceptible infections, yielding a cost-benefit ratio of 1:4. The sNB of the model is compared to two simple decision strategies: assuming that every infection is resistant (all resistant) and that no infection is resistant (all susceptible). The sNB can reach a maximum value of 1, equivalent to assuming that all resistant and susceptible cases are treated correctly (TPR=1 and FPR=0). Analyses were performed with Python 3.7, using the following packages: Numpy 1.20.3, Pandas 1.3.5 and Scikit-learn 1.0.1 for data processing; Scikit-learn, Xgboost 1.5.0, and Tensorflow 2.4.1 for modeling; Matplotlib 3.5.0 for plotting; and SHAP 0.40.0 for variable influence.

## Results

We trained four base learners, and an ensemble model composed of these base learners, to predict ciprofloxacin resistance for six bacterial species. The demographics and basic clinical characteristics corresponding to the cultures’ patients are shown in Table 1. We note that *K*.*pneumoniae* and *M*.*morganii* had a higher proportion of resistant samples in the test set, which potentially may harm predictions. Regardless, our algorithms were able to generalize successfully and achieve high ROC-AUC scores.

**Table 1:**
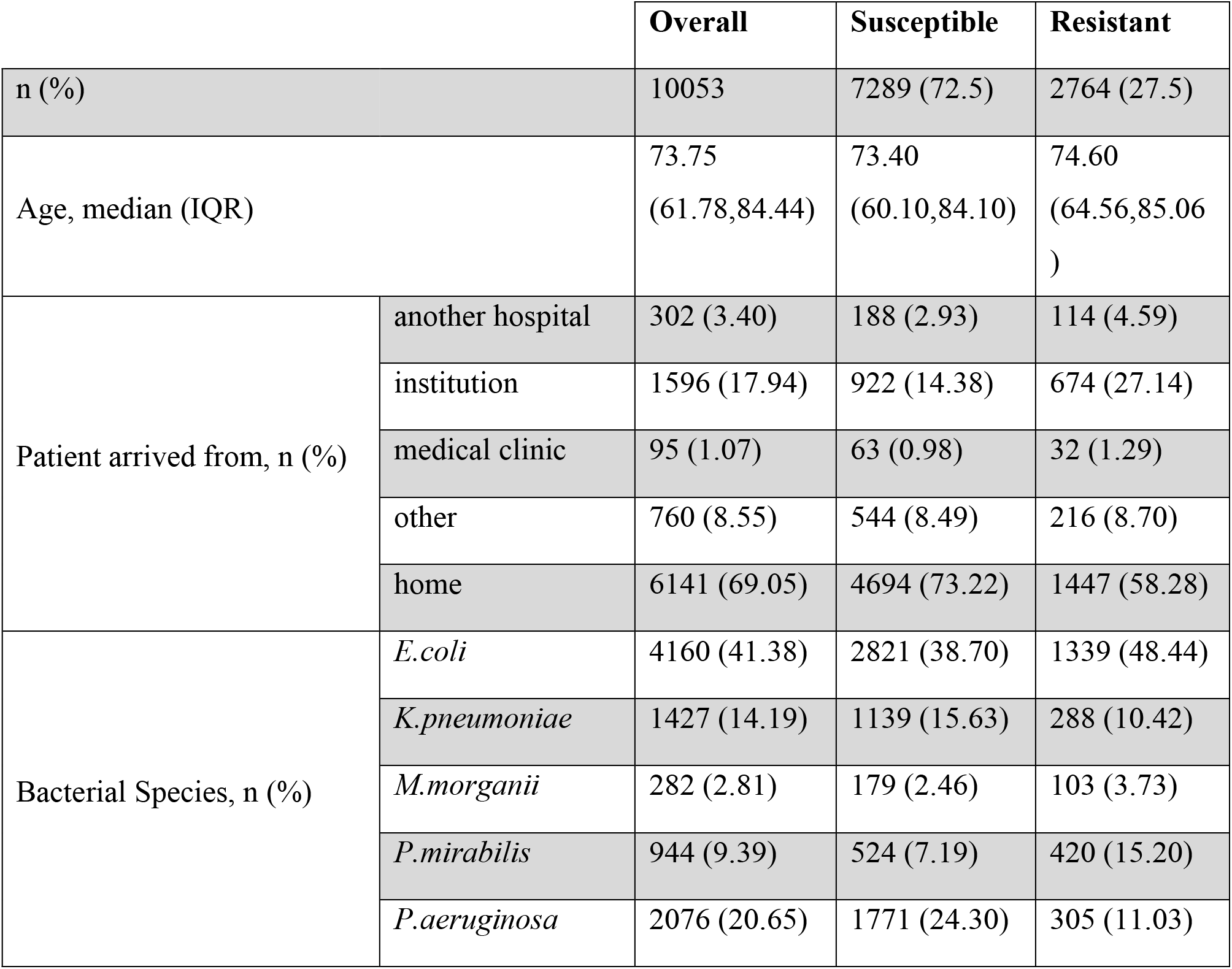
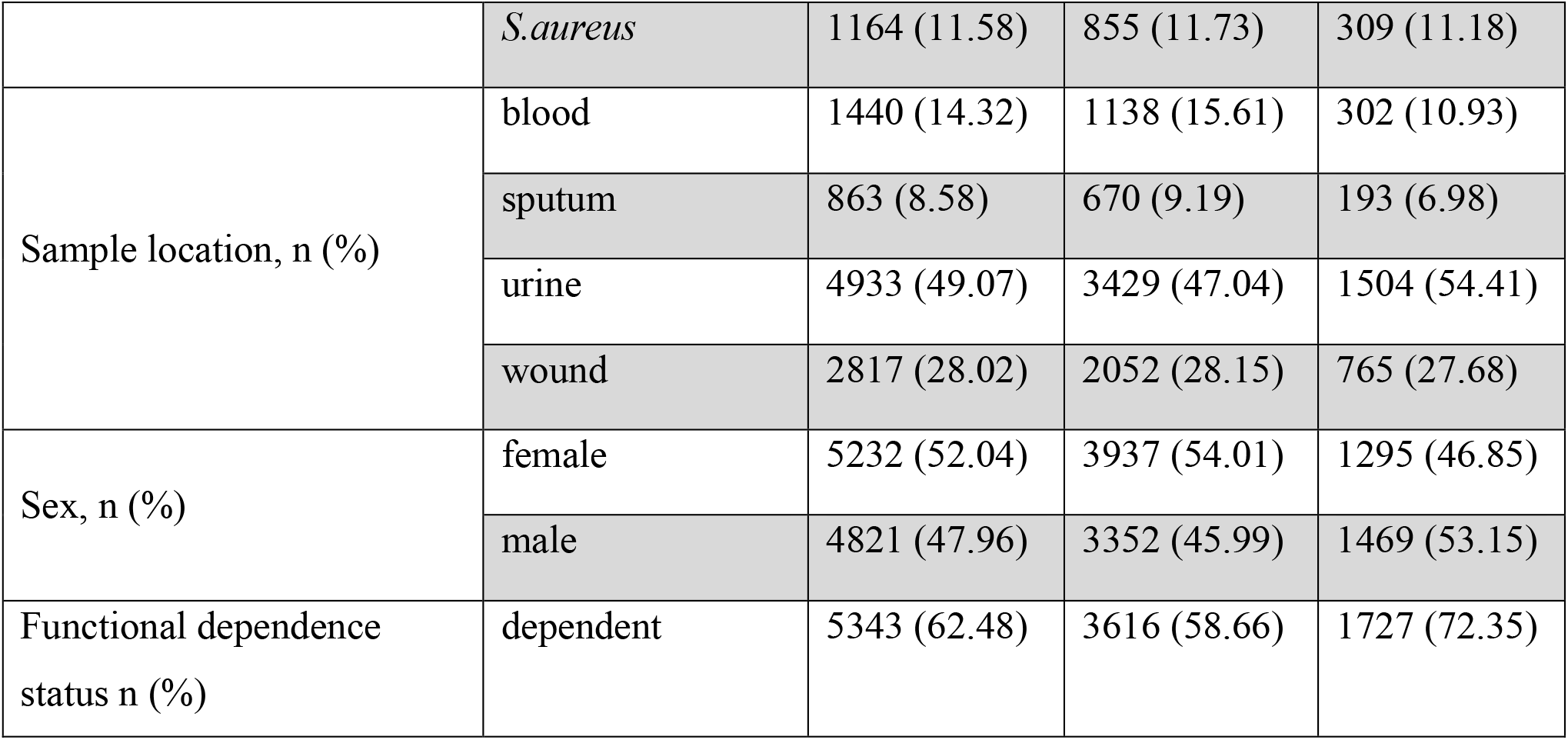
Distribution of variable values, stratified by ciprofloxacin resistance. IQR - interquartile range; SD - standard deviation.

ROC-AUC scores and calibration plots were calculated for all of the base learners (Figure 2 A-B). The ensemble consistently outperformed all base learners, on both datasets, achieving high ROC-AUC scores. For the bacteria-agnostic dataset, the ROC-AUC scores were 0.742 for the neural network, 0.739 for the logistic regression (LASSO), 0.743 for the random forest, 0.727 for the xgboost and 0.763 (95% CI 0.634-0.785) for the ensemble. On the bacteria-gnostic dataset the scores were 0.84 for the neural network, 0.84 for the LASSO, 0.823 for the random forest, 0.843 for the xgboost and 0.849 (95% CI 0.799-0.921) for the ensemble. Furthermore, all base learners and our ensemble models were relatively well-calibrated (Figure 2C-D).

**Figure 2:**
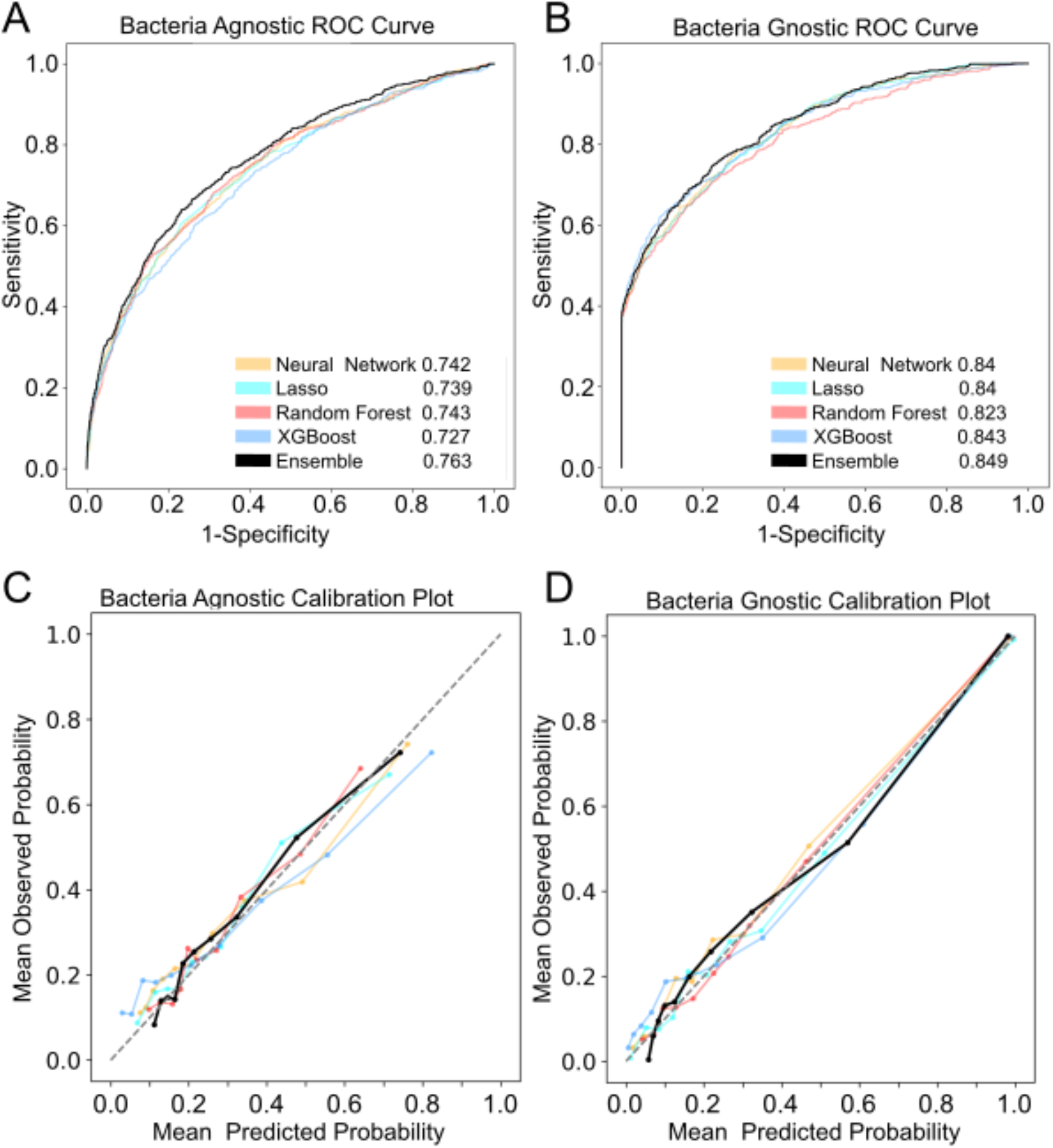
ROC curves (A, B) and calibration plots (C, D) for bacteria-agnostic and bacteria-gnostic datasets, respectively. ROC-AUC results of each model, on the test set, are presented within A and B. The colors represent different algorithms, where the black bold lines are the results of the ensemble model. Data points presented on the calibration plots are aggregated by deciles of predicted probability.

In an effort to improve the ensemble’s transparency and gain a better comprehension of the variables influencing its predictions, we used Kernel SHAP. This method estimates the contribution of each variable to the model’s prediction by approximating their SHAP values (29). These SHAP values allow us to understand the magnitude and direction of influence of variables, which implies variable importance (Figure 3).

**Figure 3:**
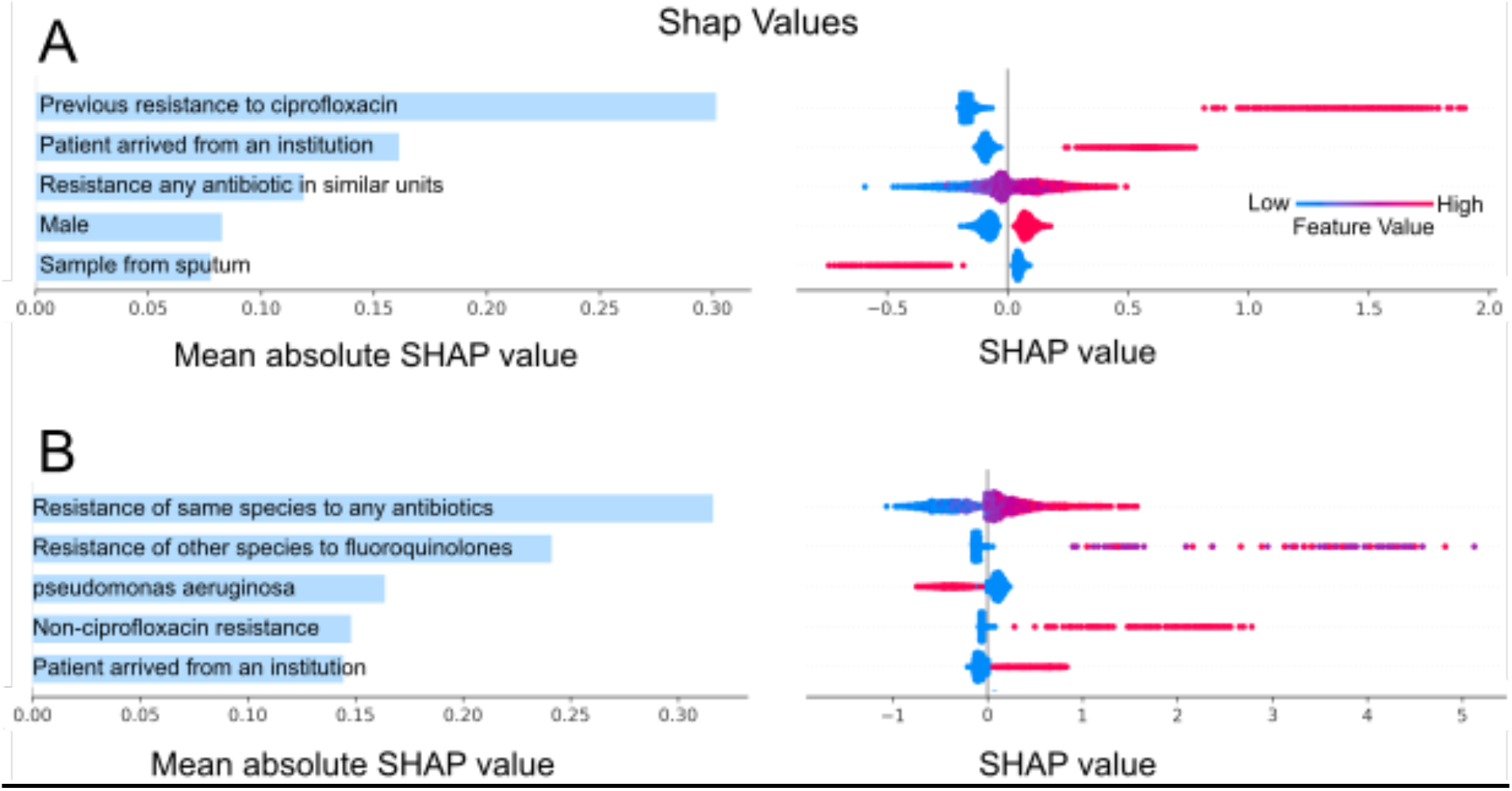
SHAP values of the ensemble model for the five most influential variables in the agnostic (A) and gnostic (B) datasets. The absolute SHAP values are presented in the left column. A swarm plot is presented in the right column, wherein colors (from blue to red) correspond to variable values (from low to high), whereas the influence of those variables on the log-OR of predictions is given on the x-axis. Previous resistance to ciprofloxacin - whether the patient had a ciprofloxacin resistant infection in the past 60 days. Resistance to any antibiotic in similar units - past 30 days moving average of resistance to any antibiotic in the same type of units (orthopedic, gynecology etc.). Resistance of the same bacterial species to any antibiotic - past 30 days moving average across the hospital. Resistance to other species to fluoroquinolones - the past 60 days, in the same patient. Non-ciprofloxacin resistance - number of non-ciprofloxacin antibiotics that the same bacterial species was resistant to in the last 60 days, in the same patient.

For the agnostic dataset, the five most influential variables in the bacteria agnostic dataset, as measured by the mean absolute SHAP values (Figure 3A), were: previous resistance to ciprofloxacin in the past 60 days, whether the patient arrived from an institution, recent resistance to any antibiotic in same type of units (e.g., internal medicine or orthopedic units), male sex, and whether the sample source was sputum. Analogously, the five most influential variables in the bacteria gnostic dataset were (Figure 3B): average resistance of the same bacterial species to any antibiotic in the past 30 days, across the hospital; the number of previous fluoroquinolone resistant infections the patient had in the past 60 days; whether the bacterial species was *P. aeruginosa*; the number of non-ciprofloxacin antibiotics that the same bacterial species had resistance to in the past 60 days, in the same patient; and whether the patient arrived from an institution. In both agnostic and gnostic settings, higher values of the influential variables consistently yielded positive influence on the ensemble’s prediction, as can be seen by the swarm plots of the SHAP values (Figure 3). This is simply the result of our coding of the binary variables’ (i.e., deciding which variable levels are set to zero or one) as risk factors.

Finally, we have performed a decision curve analysis (see Methods). Figure 4 shows that relying on predictions of our model improves assuming that every infection is resistant for cost-benefit ratios >1:9. For the gnostic model, our model always outperforms the decision rule in which we assume that every infection is susceptible (Figure 4B). However, for the agnostic model, this is true only up to a cost-benefit ratio of approximately 3:2.

**Figure 4:**
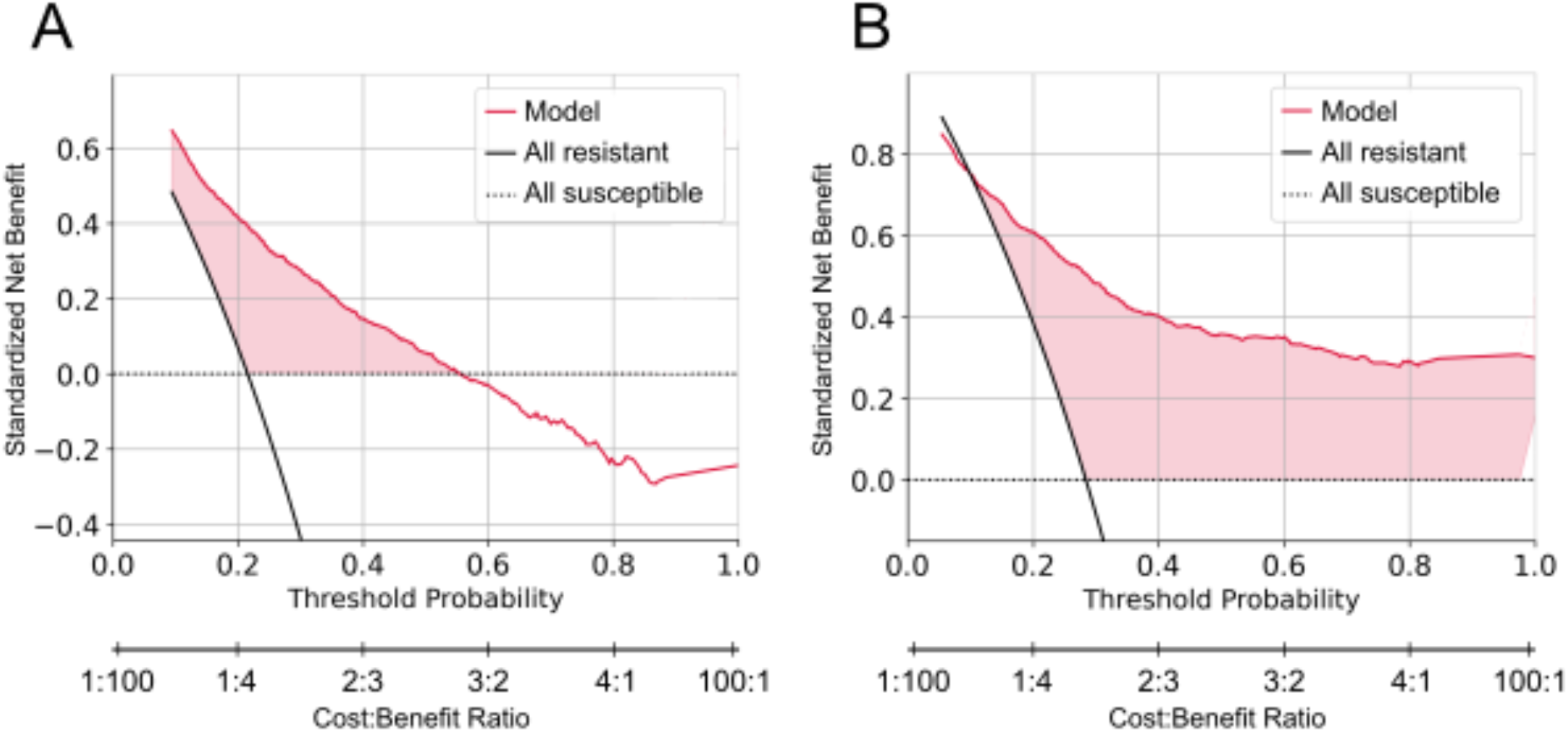
agnostic (A) and gnostic (B) decision curves. The standardized net benefit is plotted against the threshold probability and cost-benefit ratio of deciding that an infection is resistant to ciprofloxacin. Curves of the benefit when assuming all infections are susceptible (dashed horizontal line), all infections are resistant (black curve), and relying on the ensemble model predictions (red curve) are plotted. Positive differences in standardized net benefit of the model predictions vs the treat all and treat none curves are shaded in red.

## Discussion

In this study, we developed two ensemble ML models to predict resistance to ciprofloxacin of hospitalized patients’ infections. The first model was trained on the bacteria agnostic dataset, i.e., without any knowledge of the infecting bacterial species. This represents the most common situation before the start of antibiotic treatment. The second ensemble was trained on the bacteria gnostic dataset, i.e., with primary information of the infecting bacterial species. Both models achieved high ROC-AUC metrics on an independent test set: 0.763 (95% CI 0.634, 0.785) and 0.849 (95% CI 0.799, 0.921) for the agnostic and gnostic datasets, respectively, and were well calibrated. Moreover, a decision curve analysis revealed that implementing our models can be beneficial in a wide range of cost-benefit considerations of withholding vs prescribing ciprofloxacin.

Our ML models include several innovative components in the field of AMR prediction. First, we use a super learner that is trained to effectively combine the outputs of several base learners. This increases our final ROC-AUC by up to 0.036 with respect to the base-learners. Second, we incorporate variables representing recent and local resistant patterns within the hospital, in addition to a specific patient’s EMR. Consequently, and despite the limited ability to compare such results between different settings, our models achieve high predictive abilities relative to previous studies (20, 21). Importantly, our models perform well on a very heterogeneous dataset, comprising various bacterial species, sample sources and multiple departments of the hospital. For example, Feretzakis et al. predicted ciprofloxacin resistance using data from a single internal medicine department, conditioned on the sample’s Gram stain result, and reached an ROC-AUC of 0.726 (21). Yelin et al. predicted ciprofloxacin resistance only in outpatients, strictly using urine samples, and limited to three bacterial species, reaching an ROC-AUC of 0.83 (20). Other studies either did not calculate ROC-AUC (23, 32) or used cultures derived from a single sample source (22, 23), a single bacterial species (32), or a single hospital unit (33).

An additional advantage of our ensemble modeling approach is built-in model calibration. Due to the logistic transformation the single-model outputs undergo, we are able to provide an output of well-calibrated probabilities of resistance. Prescribing antibiotics forces the clinician to make a compromise between patient’s care and population-level consequences (34). Hence, providing clinicians with unbiased probabilities of resistance can facilitate incorporation of other considerations into their decision. However, we note that continuous outputs from antibiotic prescription decision-support systems have been suggested to promote over-prescription of antibiotics, and hence decisions on output forms should be made with caution (35).

Our models’ predictions were analyzed using SHAP values, which can aid in assessing the influence of different covariates on predictions when applying complex ML models (36). We note that SHAP values contain inherent flaws (37) in approximating the impact of variables on predictions, and certainly do not aim to estimate causal effects. Despite these drawbacks, SHAP values can be useful for validating model outcomes against prior knowledge of risk factors and increase models’ transparency. This can in turn facilitate increasing clinicians’ trust in using ML decision support systems in their practice (38).

The results of our SHAP analyses are indeed consistent with the literature. Highly influential variables on the ensemble models’ predictions were related to previous infections containing resistant bacteria, either to ciprofloxacin or other antibiotics. Whereas previous resistance to ciprofloxacin is an obvious risk factor for current resistance, the importance of previous resistance to other antibiotics may be explained by cross-resistance (39–41), or confounding by the patients’ exposure to resistant bacteria or to antibiotics. Patients’ origin (home, another hospital, nursing home, medical clinic, or other) had substantial influence on predictions and was also found to be an important variable by others (22, 42). This is a known risk factor, as antibiotics are administered more frequently in medical facilities and nursing homes, leading to high selection for resistance (43). Another influential variable was sex. Associations between antibiotic resistance and sex have been observed repeatedly and may stem from differential antibiotic consumption patterns (21, 22). Local resistance frequencies, which we introduced into the data as moving averages of resistance frequencies, were also found highly influential on prediction. This is consistent with previous research and clinical use of local antibiograms, representing the susceptibility patterns of different bacteria (44). Furthermore, our moving average of resistance frequencies is potentially more sensitive to resistance trends than yearly or monthly antibiograms. In the gnostic model, *P. aeruginosa* was selected as an influential variable. This stems from the binary encoding of the bacterial species, which defined the reference species as *E. coli*. Since *P. aeruginosa* was the second-most common bacterial species in the dataset, and was less resistant than *E. coli* (Table 1), it was determined to be influential in reducing the predicted probability of a resistant infection.

Our study has several limitations. First, our dataset lacks relevant community-related patient information, such as antibiotic consumption in the community (43), and antibiotic consumption in the patients’ surroundings, including neighborhoods (15) and households (45). Our models can be easily extended to accommodate these covariates, which will likely further improve the models’ predictive abilities. Second, our models are not necessarily immediately generalizable to other settings, or even the same setting, in different time periods. Variations in antibiotic consumption and the dynamic nature of AMR may lead to variation in risk factors over space and time (46, 47). Retraining of the models on site-specific data will likely be required to fine-tune predictions in different settings. However, the rates of ciprofloxacin resistance and patient covariates in our dataset are comparable with those of other developed countries (48). We therefore expect a reasonable degree of consistency in our results, if our models would have been developed on a dataset from comparable settings.

## Conclusions

The models developed in this study represent a further step on the way to inclusion of ML decision support systems into clinical practice. Improvement of such models depends on advances in algorithm development, specific feature engineering, and the augmentation of the quantity and quality of EMR data. As we have shown, modern ML models can achieve high prediction while autonomously imparting high influence to risk factors that are known to be clinically relevant to AMR. Hopefully, future studies can further leverage the presented models and the vast EMR data available to improve prediction of AMR and consequently reduce antibiotic misuse.

## Data Availability

Data are proprietary but can be made available upon reasonable request from the authors.

## Declarations

### Ethics approval

The study was approved by the Institutional Review Board (Helsinki) Committee of Meir Medical Center. Since this was a retrospective study, using archived medical records, an exemption from informed consent was granted by the Helsinki Committee.

### Data availability

Data are proprietary but can be made available upon reasonable request from the authors.

### Transparency declarations

None to declare.

### Author contributions

IM and UO conceived the study; IM implemented the analysis; IM, MC and UO interpreted the results; IM and UO wrote the initial draft of the manuscript; all authors revised and approved the final version of the manuscript.

## References

1. Smith RA, M’ikanatha NM, Read AF. 2015. Antibiotic resistance: a primer and call to action. Health Commun 30:309–314.

2. Palumbi SR. 2001. Humans as the world’s greatest evolutionary force. Science 293:1786–1790.

3. Weber DJ. 2006. Collateral damage and what the future might hold. The need to balance prudent antibiotic utilization and stewardship with effective patient management. Int J Infect Dis 10:S17–S24.

4. Carrara E, Pfeffer I, Zusman O, Leibovici L, Paul M. 2018. Determinants of inappropriate empirical antibiotic treatment: systematic review and meta-analysis. Int J Antimicrob Agents 51:548–553.

5. Organization WH. 2019. Executive summary: the selection and use of essential medicines 2019: report of the 22nd WHO Expert Committee on the selection and use of essential medicines: WHO Headquarters, Geneva, 1-5 April 2019. World Health Organization.

6. Chowers M, Zehavi T, Gottesman B-S, Baraz A, Nevo D, Obolski U. 2022. Estimating the impact of cefuroxime versus cefazolin and amoxicillin/clavulanate use on future collateral resistance: a retrospective comparison. J Antimicrob Chemother.

7. Nathwani D, Varghese D, Stephens J, Ansari W, Martin S, Charbonneau C. 2019. Value of hospital antimicrobial stewardship programs [ASPs]: a systematic review. Antimicrob Resist Infect Control 8:35.

8. Tribble AC, Lee BR, Flett KB, Handy LK, Gerber JS, Hersh AL, Kronman MP, Terrill CM, Sharland M, Newland JG. 2020. Appropriateness of antibiotic prescribing in United States children’s hospitals: a national point prevalence survey. Clin Infect Dis 71:e226–e234.

9. eEML - Electronic Essential Medicines List. https://list.essentialmeds.org/. Retrieved 10 July 2022.

10. Loscalzo J, Fauci AS, Kasper DL, Hauser S, Longo D, Jameson JL. 2022. Harrison’s Principles of Internal Medicine, (Vol. 1 & Vol. 2). McGraw Hill Professional.

11. Sharma PC, Jain A, Jain S, Pahwa R, Yar MS. 2010. Ciprofloxacin: review on developments in synthetic, analytical, and medicinal aspects. J Enzyme Inhib Med Chem 25:577–589.

12. Thomson CJ. 1999. The global epidemiology of resistance to ciprofloxacin and the changing nature of antibiotic resistance: a 10 year perspective. J Antimicrob Chemother 43:31–40.

13. World Health Organization. 2021. Global antimicrobial resistance and use surveillance system (GLASS) report: 2021.

14. Dalhoff A. 2012. Global Fluoroquinolone Resistance Epidemiology and Implictions for Clinical Use. Interdiscip Perspect Infect Dis 2012:e976273.

15. Low M, Neuberger A, Hooton TM, Green MS, Raz R, Balicer RD, Almog R. 2019. Association between urinary community-acquired fluoroquinolone-resistant Escherichia coli and neighbourhood antibiotic consumption: a population-based case-control study. Lancet Infect Dis 19:419–428.

16. Eliopoulos GM, Cosgrove SE, Carmeli Y. 2003. The Impact of Antimicrobial Resistance on Health and Economic Outcomes. Clin Infect Dis 36:1433–1437.

17. Gottesman BS, Carmeli Y, Shitrit P, Chowers M. 2009. Impact of quinolone restriction on resistance patterns of Escherichia coli isolated from urine by culture in a community setting. Clin Infect Dis 49:869–875.

18. Anahtar MN, Yang JH, Kanjilal S. 2021. Applications of Machine Learning to the Problem of Antimicrobial Resistance: an Emerging Model for Translational Research. J Clin Microbiol 59:e01260–20.

19. Rawson TM, Ahmad R, Toumazou C, Georgiou P, Holmes AH. 2019. Artificial intelligence can improve decision-making in infection management. 6. Nat Hum Behav 3:543–545.

20. Yelin I, Snitser O, Novich G, Katz R, Tal O, Parizade M, Chodick G, Koren G, Shalev V, Kishony R. 2019. Personal clinical history predicts antibiotic resistance of urinary tract infections. 7. Nat Med 25:1143–1152.

21. Feretzakis G, Loupelis E, Sakagianni A, Kalles D, Martsoukou M, Lada M, Skarmoutsou N, Christopoulos C, Valakis K, Velentza A, Petropoulou S, Michelidou S, Alexiou K. 2020. Using Machine Learning Techniques to Aid Empirical Antibiotic Therapy Decisions in the Intensive Care Unit of a General Hospital in Greece. 2. Antibiotics 9:50.

22. Dan S, Shah A, Justo JA, Bookstaver PB, Kohn J, Albrecht H, Al-Hasan MN. 2016. Prediction of Fluoroquinolone Resistance in Gram-Negative Bacteria Causing Bloodstream Infections. Antimicrob Agents Chemother 60:2265–2272.

23. Dickstein Y, Geffen Y, Andreassen S, Leibovici L, Paul M. 2016. Predicting Antibiotic Resistance in Urinary Tract Infection Patients with Prior Urine Cultures. Antimicrob Agents Chemother 60:4717–4721.

24. Eilers PH, Boer JM, van Ommen G-J, van Houwelingen HC. 2001. Classification of microarray data with penalized logistic regression, p. 187–198. In Microarrays: optical technologies and informatics. International Society for Optics and Photonics.

25. Hastie T, Tibshirani R, Friedman JH, Friedman JH. 2009. The elements of statistical learning: data mining, inference, and prediction. Springer.

26. Bergstra J, Bengio Y. 2012. Random search for hyper-parameter optimization. J Mach Learn Res 13.

27. Sill J, Takacs G, Mackey L, Lin D. 2009. Feature-Weighted Linear Stacking. ArXiv09110460 Cs.

28. Van der Laan MJ, Polley EC, Hubbard AE. 2007. Super learner. Stat Appl Genet Mol Biol 6.

29. Lundberg SM, Lee S-I. A Unified Approach to Interpreting Model Predictions 10.

30. Vickers AJ, Elkin EB. 2006. Decision curve analysis: a novel method for evaluating prediction models. Med Decis Making 26:565–574.

31. Kerr KF, Brown MD, Zhu K, Janes H. 2016. Assessing the Clinical Impact of Risk Prediction Models With Decision Curves: Guidance for Correct Interpretation and Appropriate Use. J Clin Oncol Off J Am Soc Clin Oncol 34:2534–2540.

32. Gallini A, Degris E, Desplas M, Bourrel R, Archambaud M, Montastruc J-L, Lapeyre-Mestre M, Sommet A. 2010. Influence of fluoroquinolone consumption in inpatients and outpatients on ciprofloxacin-resistant Escherichia coli in a university hospital. J Antimicrob Chemother 65:2650–2657.

33. Wang T, Hansen KR, Loving J, Paschalidis IC, van Aggelen H, Simhon E. 2021. Predicting Antimicrobial Resistance in the Intensive Care Unit. 2111.03575. arXiv.

34. Wojcik G, Ring N, McCulloch C, Willis DS, Williams B, Kydonaki K. 2021. Understanding the complexities of antibiotic prescribing behaviour in acute hospitals: a systematic review and meta-ethnography. Arch Public Health 79:134.

35. Diamant M, Baruch S, Kassem E, Muhsen K, Samet D, Leshno M, Obolski U. 2021. A game theoretic approach reveals that discretizing clinical information can reduce antibiotic misuse. Nat Commun 12:1–13.

36. Shapley LS. 1953. A value for n-person games, Contributions to the theory of games II (AW Tucker and HW Kuhn, eds.). Princeton University Press, Princeton, USA.

37. Kumar IE, Venkatasubramanian S, Scheidegger C, Friedler S. 2020. Problems with Shapley-value-based explanations as feature importance measures, p. 5491–5500. In International Conference on Machine Learning. PMLR.

38. Chen M, Zhang B, Cai Z, Seery S, Mendez MJ, Ali NM, Ren R, Qiao Y-L, Xue P, Jiang Y. 2022. Physician and Medical Student Attitudes Toward Clinical Artificial Intelligence: A Systematic Review with Cross-Sectional Survey. 4128867. SSRN Scholarly Paper. Rochester, NY https://doi.org/10.2139/ssrn.4128867.

39. Cherny SS, Nevo D, Baraz A, Baruch S, Lewin-Epstein O, Stein GY, Obolski U. 2021. Revealing antibiotic cross-resistance patterns in hospitalized patients through Bayesian network modelling. J Antimicrob Chemother 76:239–248.

40. Cherny SS, Chowers M, Obolski U. 2022. Patterns of antibiotic cross-resistance by bacterial sample source: a retrospective cohort study. medRxiv.

41. Beckley AM, Wright ES. 2021. Identification of antibiotic pairs that evade concurrent resistance via a retrospective analysis of antimicrobial susceptibility test results. Lancet Microbe 2:e545–e554.

42. Lewin-Epstein O, Baruch S, Hadany L, Stein GY, Obolski U. 2021. Predicting antibiotic resistance in hospitalized patients by applying machine learning to electronic medical records. Clin Infect Dis 72:e848–e855.

43. Chatterjee A, Modarai M, Naylor NR, Boyd SE, Atun R, Barlow J, Holmes AH, Johnson A, Robotham JV. 2018. Quantifying drivers of antibiotic resistance in humans: a systematic review. Lancet Infect Dis 18:e368–e378.

44. Truong WR, Hidayat L, Bolaris MA, Nguyen L, Yamaki J. 2021. The antibiogram: Key considerations for its development and utilization. JAC-Antimicrob Resist 3:dlab060.

45. Oonsivilai M, Mo Y, Luangasanatip N, Lubell Y, Miliya T, Tan P, Loeuk L, Turner P, Cooper BS. 2018. Using machine learning to guide targeted and locally-tailored empiric antibiotic prescribing in a children’s hospital in Cambodia. Wellcome Open Res 3:131.

46. Bell BG, Schellevis F, Stobberingh E, Goossens H, Pringle M. 2014. A systematic review and meta-analysis of the effects of antibiotic consumption on antibiotic resistance. BMC Infect Dis 14:1–25.

47. Baraz A, Chowers M, Nevo D, Obolski U. 2022. Stable temporal relationships as a first step towards causal inference: an application to antibiotic resistance. preprint. Epidemiology.

48. Fasugba O, Gardner A, Mitchell BG, Mnatzaganian G. 2015. Ciprofloxacin resistance in community- and hospital-acquired Escherichia coli urinary tract infections: a systematic review and meta-analysis of observational studies. BMC Infect Dis 15:545.

