## Supplementary Material for "Prediction of Ciprofloxacin Resistance in Hospitalized Patients Using Machine Learning"

**Figure S1:** An illustration of the ensemble model pipeline (1). L1-LASSO, RF-random forest, XGB-xgboost, NN-neural network, Y-true resistance

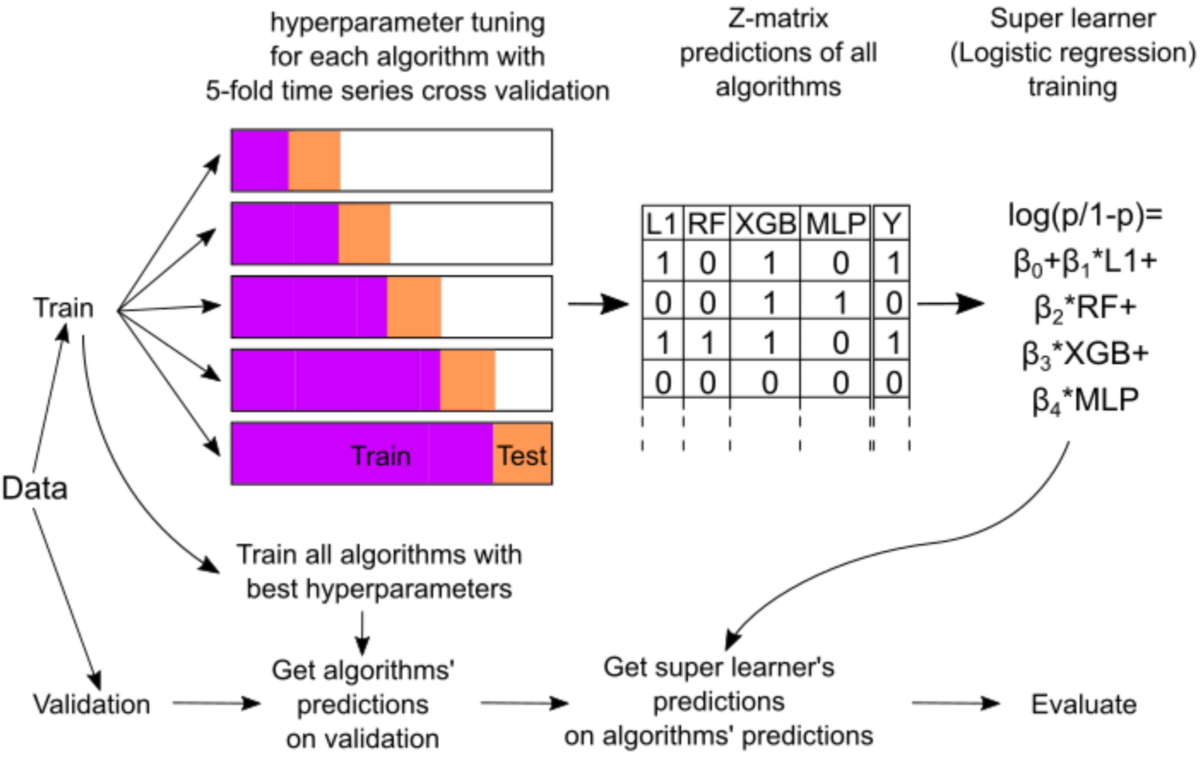


**Table S1:** Features that were used and their meaning

| Bacteria agnostic variables | |
|---|---|
| **Name** | **Meaning** |
| Age.At.Culture.Time.Date | Age |
| Arrived.From | The place from which the patient arrived to a general unit in the hospital (their home, institution, another hospital, medical clinic, other) |
| COPD | Whether the patient has chronic obstructive pulmonary disease |
| CRF | Whether the patient has chronic heart failure |

| | |
|---|---|
| Charlson.Score | A comorbidity based score. Based on the ICD9 codes in the admission data and the Charlson comorbidity system. In case of no known comorbidities, this score was assigned 0. |
| Chronic.Wound | Whether the patient has chronic wound |
| Culture.Year | The year from cutlture date |
| CumAnyUsage365 | Cumulative usage of any antibiotics in the last year since current culture date |
| Dementia | Whether the patient has dementia |
| Diabetes | Whether the patient has diabetes |
| Immunosuppression | Whether the patient ever suffered from an immunosuppressive disease prior to the time when the resistance culture was taken |
| Invasive.Procedure | Whether the patient underwent an invasive procedure prior to the time when the resistance culture was taken |
| Is.ER | Whether the culture was taken in the ER |
| Is.Meds.Sensitive | Whether the patient is sensitive to any medication |
| Is.Nosocomial | Whether the patient was in the hospital 48 hours prior to the susceptibility test, because then there's a good chance that the infection is nosocomial. |
| Length.Days.Current.Hospitalization | Length (in days) of current hospitalization (up to the culture's time and date) |
| Length.Days.Hospitalized.Past.365.Days | Number of days hospitalized in the 365 days before the Culture's Time & Date, excluding the current hospitalization |

| | |
|---|---|
| Obesity | Whether the patient is obese |
| Original.Unit | internal d, internal b, orthopedics a, internal c, surgery a, geriatric rehab, internal e, lungs, internal a, maternity, orthopedics b ,surgery b, internal ER , surgery ER, internal f, orthopedic ER, urology, pediatric, vascular surgery, plastic surgery, spine surgery, women, maternity ward b, chest surgery, ENT, pediatric emergency medicine, pediatric surgery, internal icu - respiratory, pediatric orthopedics, Oral and maxillofacial surgery, mother and fetus, pediatric plastic surgery, maternity ward c, neonatal, pediatric urology, walkin ER, women ER |
| PrevAntiUsageBin60<br>PrevAntiUsageBin61_180<br>PrevAntiUsageBin181_365 | Cumulative usage (in days) of the ciprofloxacin in the previous (0-60/61-180/181-365) days. Excluding the day before the culture was taken. |
| PrevAntiUsageFamilyBin60<br>PrevAntiUsageFamilyBin61_180<br>PrevAntiUsageFamilyBin181_365 | Cumulative usage (in days) of fluoroquinolone in the previous (0-60/61-180/181-365) days. Excluding the day before the culture was taken. |
| PrevAntiUsageOtherBin60<br>PrevAntiUsageOtherBin61_180<br>PrevAntiUsageOtherBin181_365 | Cumulative usage (in days) of other than ciprofloxacin in the previous (0-60/61-180/181-365) days. Excluding the day before the culture was taken. |
| PrevAntiUsageOtherFamilyBin60<br>PrevAntiUsageOtherFamilyBin61_180<br>PrevAntiUsageOtherFamilyBin181_365 | Cumulative usage (in days) of other than fluoroquinolone in the previous (0-60/61-180/181-365) days. Excluding the day before the culture was taken. |

| | |
|---|---|
| PreviouslyResistantOtherAntiSameFamilyBin60<br>PreviouslyResistantOtherAntiSameFamilyBin61_181<br>PreviouslyResistantOtherAntiSameFamilyBin181_365 | Boolean - True if the patient had resistance to fluoroquinolone (excluding ciprofloxacin) during the previous (0-60/61-180/181-365) days (from culture time date) |
| PreviouslyResistantOtherFamilyBin60<br>PreviouslyResistantOtherFamilyBin61_180<br>PreviouslyResistantOtherFamilyBin181_365 | Boolean - True if the patient had resistance to non fluoroquinolone during the previous (0-60/61-180/181-365) days (from culture time date) |
| PreviouslyResistantSameAnti | Weather the last time (ever) the patient was hospitalized there was resistance to the tested antibiotics |
| PreviouslyResistantSameAntiBin60<br>PreviouslyResistantSameAntiBin61_180<br>PreviouslyResistantSameAntiBin180_365 | Boolean - True if the patient had resistance to the ciprofloxacin during the previous (0-60/61-180/181-365) days (from culture time date) |
| Sample.Location | Where was the culture was taken (blood, sputum, urine, wound) |
| Sex | Patients sex |
| SlidingResAny30 | Rolling mean of last 30 days of resistance to any |
| SlidingResSame30 | Rolling mean of last 30 days of resistance to this antibiotic |
| SlidingResUnits30 | Rolling mean of resistance of the last 30 days in similar units (internals orthopedics ERs urgeries) to any antibiotic |
| Status.Before | The medical status of the patient before arriving at the hospital (independent, dependent) |
| TimeSinceEndLastUse365 | Time in days since the last use of ciprofloxacin during the last year |
| Times.Hospitalized.Past.365.Days | Number of times hospitalized in the |

| | |
|---|---|
| | 365 days before the Culture's Time & Date, excluding the current hospitalization |
| Unit | Unit of hospitalization (internal, orthopedic, surgical, geriatric, obstetrics, ER, children, gynecology, pediatric surgery, ICU) |
| UnqAntibiotics365 | Number of unique antibiotics that were used during the last year since current culture date |
| UnqFamilies365 | Number of unique antibiotic families that were used during the last year since current culture date |
| Van.Walraven.Score | A comorbidity based score. Based on the ICD9 codes in the admission data and the Elixhauser comorbidity system. In case of no known comorbidities, this score was assigned 0 |
| **Bacteria gnostic additional variables** | |
| Bacterium | Escherichia coli, Klebsiella pneumoniae, Morganella morganii, Proteus mirabilis, Pseudomonas aeruginosa, Staphylococcus aureus |
| BoolSameBacSameAntiResBin60<br>BoolSameBacSameAntiResBin61_365<br>BoolSameBacSameAntiResBin181_365 | Boolean - True if the same bacterium had resistance to the ciprofloxacin in the previous (0-60/61-180/181-365) days.Excluding the day before the culture was taken. |
| BoolSameBacSameFamilyResBin60<br>BoolSameBacSameFamilyResBin61_180<br>BoolSameBacSameFamilyResBin181-365 | Boolean - True if the same bacterium had resistance to fluoroquinolone in the previous (0-60/61-180/181-365) days. Excluding the day before the culture was taken. |

| | |
|---|---|
| PrevOtherBacSameFamilyResBin60<br>PrevOtherBacSameFamilyResBin61_180<br>PrevOtherBacSameFamilyResBin181_365 | The number of times other than tested bacteria that had resistance to fluoroquinolone antibiotic during (0-60/61-180/181-365) days prior to the current culture. |
| SlidingResSameBacAnyAnti30 | Rolling mean of last 30 days of the same bacterium for the same antibiotic |
| UnqOtherBacSameAntiResBin60<br>UnqOtherBacSameAntiResBin61_180<br>UnqOtherBacSameAntiResBin181_365 | The number of unique bacteria that had resistance to the same as tested antibiotic during (0-60/61-180/181-365) days prior to the current culture. |
| UnqSameBacOtherAntiResBin60<br>UnqSameBacOtherAntiResBin61_180<br>UnqSameBacOtherAntiResBin181_365 | The number of unique antibiotics other than ciprofloxacin that the same bacterium had resistance to in the previous (0-60/61-180/181-365) days (from culture time date) |
| UnqSameBacOtherFamilyResBin60<br>UnqSameBacOtherFamilyResBin61_180<br>UnqSameBacOtherFamilyResBin181_365 | The number of unique antibiotics other than fluoroquinolone that the same bacterium had resistance to in the previous (0-60/61-180/181-365) days (from culture time date) |

**Table S2:** Hyperparameters of the final models. Showing only non-default values.

| Algorithm | Hyperparameter | Agnostic dataset | Gnostic dataset |
|---|---|---|---|
| Neural Network | activation | linear | sigmoid |
| | batch_size | 68 | 172 |
| | beta_1 | 0.8909636176119624 | 0.28377762986855604 |
| | beta_2 | 0.7328189557393326 | 0.7508823267588157 |
| | dropout_rate | 0.38126629372786847 | 0.28377762986855604 |
| | epochs | 12 | 13 |

| | | | |
|---|---|---|---|
| | learn_rate | 0.0022839151250675584 | 0.003990228421227295 |
| | n_layers | 1 | 1 |
| | neurons1 | 84 | 31 |
| | neurons2 | 42 | 16 |
| | optimizer | Adam | Adam |
| **Logistic regression** | C | 0.2686236831651222 | 0.861569882192526 |
| | max_iter | 10000 | 10000 |
| | penalty | l1 | l1 |
| | solver | saga | saga |
| **Random Forest** | bootstrap | False | False |
| | max_depth | 13 | 14 |
| | max_features | 0.4740361409628354 | 0.4856061544026114 |
| | min_samples_split | 0.049136994172804475 | 0.038244056567505284 |
| | n_estimators | 466 | 326 |
| **XGBoost** | colsample_bytree | 0.9991525259766447 | 0.9332786246730219 |
| | gamma | 2.772874372213767 | 2.0437254708124595 |
| | learning_rate | 0.5123765489387977 | 0.2688917726724662 |
| | max_depth | 13 | 5 |
| | min_child_weight | 7.518775429130775 | 1.934384043661038 |
| | n_estimators | 325 | 302 |
| | subsample | 0.9644823255038443 | 0.9567242515147112 |
| **Ensemble (Logistic Regression)** | C | 0.5870235632684522 | 0.3693543790751912 |
| | max_iter | 100 | 100 |
| | penalty | l1 | l1 |

| | | | |
|---|---|---|---|
| | solver | liblinear | liblinear |
| **Ensemble coefficient** | NN | 1.4363707834813666 | 3.1721876362839816 |
| | l1 | 0.2738753340149202 | 1.498039073645653 |
| | RF | 1.9078339179490795 | 1.5192861627065386 |
| | XGBoost | 0.8743269280774444 | 0.8907533051507616 |

### Supplementary reference


1. Van der Laan MJ, Polley EC, Hubbard AE. 2007. Super learner. Stat Appl Genet Mol Biol 6.
